# Cardiorespiratory fitness indicators and Life’s Essential 8 in individuals with coronary artery disease

**DOI:** 10.64898/2026.07.06.26357366

**Authors:** Emilio J. Barranco-Moreno, Sol Vidal-Almela, Lucía Sánchez-Aranda, Anna Carlén, Marcos Olvera-Rojas, Rosa M. Alonso-Cuenca, Patricio Solis-Urra, Javier Sanchez-Martinez, Javier Fernández-Ortega, Esmee A. Bakker, Ángel Herraiz-Adillo, Pontus Henriksson, Eduardo Moreno-Escobar, Rocío García-Orta, Irene Esteban-Cornejo, Angel Toval, Francisco B. Ortega

**Affiliations:** Department of Physical and Sports Education, Sport and Health University Research Institute (iMUDS), Faculty of Sport Sciences, University of Granada, Granada, Spain; Department of Clinical Physiology in Linköping, and Department of Health, Medicine and Caring Sciences, Linköping University, Linköping, Sweden; Cardiology Service, San Cecilio Clinical University Hospital, Granada, Spain; AdventHealth Research Institute, Neuroscience Institute, Orlando, FL, USA; Faculty of Education and Social Sciences, Universidad Andres Bello, Viña del Mar 2531015, Chile; Department of Primary and Community Care, Radboud University Medical Center, Nijmegen, Netherlands; Department of Health, Medicine and Caring Sciences, Linköping University, Linköping, Sweden; Cardiology Service, Virgen de Las Nieves University Hospital, Granada, Spain; CIBER de Fisiopatología de la Obesidad y Nutrición (CIBEROBN), Instituto de Salud Carlos III, Madrid, Spain; Instituto de Investigación Biosanitaria, ibs.Granada, 18012, Granada, Spain

**Author notes:** Reprint and correspondence: Professor Francisco B. Ortega and Mr. Emilio J. Barranco-Moreno, Department of Physical and Sports Education, Faculty of Sport Sciences, University of Granada, Granada, Spain. E-mail address and.

## Abstract

**Objective:** To analyze the associations between cardiorespiratory fitness (CRF) indicators and cardiovascular health, assessed through the Life’s Essential 8 (LE8) score, in individuals with coronary artery disease (CAD).

**Patients and methods:** This cross-sectional study included individuals (aged 50-75 years) with stable CAD were enrolled in the Heart-Brain randomized controlled trial (NCT06214624) from April 2022 to June 2024. Participants underwent a cardiopulmonary exercise test until volitional exhaustion. CRF indicators included peak oxygen consumption (VO_2peak_), time to exhaustion (TTE), ventilatory anaerobic threshold (VAT), peak oxygen pulse, 60-s heart rate recovery (HRrec) and oxygen uptake efficiency slope (OUES). LE8 score (range 0-100) was calculated as the unweighted average of 8 variables: physical activity, sleep, diet, nicotine exposure, glucose, lipids, body mass index, and blood pressure, as defined by the American Heart Association. We used linear regression models adjusted for sex, age, and education.

**Results:** 102 individuals were included (21 females). VO_2peak_ (β_std_=0.67, *P*<.001) and TTE (β_std_=0.64, *P*<.001) showed strong positive associations with LE8 total score, followed by VAT (β_std_=0.43, *P*<.001) and HRrec (β_std_=0.26, *P*=.01). No associations were found for OUES (β_std_=-0.08, *P*=.54) and peak oxygen pulse (β_std_=-0.05, *P*=.72).

**Conclusion:** Maximal and submaximal indicators of CRF were positively associated with LE8 in individuals with CAD, yet maximal indicators showed the strongest associations. Notably, TTE demonstrated a similar strength of association with LE8 as VO_2peak_. These findings have important clinical and research implications as they support TTE as a simpler (than the gold-standard VO_2peak_), yet informative, marker of cardiovascular health in individuals with CAD.

**Clinical trials registration number:** NCT06214624, https://clinicaltrials.gov/study/NCT06214624

**Abstract presentation:** Preliminary results of this study were previously presented as an abstract at the “XI EXERNET Symposium (Salud y bienestar a través del ejercicio: abordajes innovadores en la Red EXERNET), Seville, Spain, October 24–25, 2025”.

## INTRODUCTION

Coronary artery disease (CAD) remains the leading cause of disability^1^ and mortality^2^ worldwide, and its burden attributable to risk factors continues to increase.^3^ Importantly, among individuals with established CAD, a substantial proportion of residual risk remains attributable to modifiable factors. A prospective cohort study estimated that modifiable risk factors account for 66% of the risk of recurrent major cardiovascular events in cardiovascular disease populations.^4^ In line, the Global Burden of Disease Study reported declines in CAD deaths in regions with better-controlled metabolic risk,^5^ suggesting that optimizing risk factor management offers potential to diminish the global burden of CAD.

In 2022, the AHA defined ideal cardiovascular health (CVH) according to the Life’s Essential 8 (LE8) score, aiming to enhance its monitoring and promotion by focusing on major modifiable risk factors.^6^ LE8 is summarized as a 0-100 score based on both health behaviors and factors.^6^ Higher LE8 scores have shown associations with lower subclinical coronary atherosclerosis^7^ and risk of all-cause and cardiovascular disease mortality in the general population,^8–12^ as well as fewer recurrent cardiovascular events^13^ and a lower mortality risk in individuals with cardiovascular disease.^11,14,15^ Therefore, LE8 is emerging as a useful measure of modifiable CVH in the secondary prevention setting in CAD, while additional objective markers may further refine risk characterization.

Cardiorespiratory fitness (CRF) is a robust predictor of morbidity and mortality among healthy and clinical populations,^16,17^ and is highly linked to CAD prognosis.^18–22^ The gold-standard method for the assessment of CRF is cardiopulmonary exercise testing (CPET), which offers objective measures from gas exchange analysis. The key physiological indicator of CRF is peak oxygen uptake (VO_2peak_). However, focusing exclusively on VO_2peak_ may limit the breadth of information captured from CPET, as other maximal and submaximal indicators have also shown prognostic capacity and allow for a more comprehensive evaluation of the cardiopulmonary response to exercise.^23–28^

Besides its prognostic value, CRF is closely associated with established CAD risk factors, with higher CRF often corresponding to a healthier risk factor profile.^29,30^ Nevertheless, the evidence linking CPET-derived CRF indicators to composite CVH scores such as LE8 remains limited. The scarce literature suggests that low CRF is associated with higher odds of poor CVH scores.^31–34^ To our knowledge, only one study has assessed the association of LE8 and CRF using CPET.^34^ Ravichandran et al. found that LE8 was favorably associated with VO_2peak_, ventilatory efficiency, resting heart rate, and blood pressure responses to exercise in a community-based sample, with consistent LE8-VO_2peak_ associations observed among participants with prevalent cardiovascular disease.^34^ However, CAD-specific evidence remains underexplored, particularly regarding a broader set of CRF indicators.

Identifying which CPET-derived indicators are more closely related to LE8 may help determine whether simpler and more feasible exercise markers can provide clinically useful information on CVH in individuals with CAD. Therefore, this study aimed to examine the associations between a comprehensive set of CRF indicators (i.e., VO_2peak_, time to exhaustion [TTE], ventilatory anaerobic threshold [VAT], oxygen uptake efficiency slope [OUES], peak oxygen pulse, and heart rate recovery after 60 seconds [HRrec]) and the LE8 score in individuals with CAD.

## METHODS

### Study design and participants

This cross-sectional analysis used baseline data from the Heart-Brain randomized controlled trial (NCT06214624; https://clinicaltrials.gov/study/NCT06214624). The trial was approved by the Research Ethics Board of the Andalusian Health Service (CEIM/CEI Provincial de Granada; #1776-N-21 on December 21st, 2021) and followed the principles of the Declaration of Helsinki. All participants provided written informed consent. Data was acquired between April 2022 and June 2024. A detailed description of the study protocol has been published elsewhere.^35^ Briefly, the Heart-Brain trial included participants aged 50-75 years with stable CAD, enrolled from two hospitals in Granada, Spain. Potentially eligible participants were identified through cardiology consultations and hospital patient lists. CAD diagnosis was confirmed by invasive coronary angiography or computed tomography showing at least one coronary stenosis with ≥50% luminal narrowing; with left ventricular ejection fraction ≥45%. Exclusion criteria included acute coronary syndrome in the last year, coronary artery bypass surgery or percutaneous coronary intervention in the last 6 months, grade III obesity (body mass index [BMI] >40 kg/m^2^), and being physically active. The Heart-Brain trial sample size was calculated to detect a low-to-medium effect on its primary outcome (cerebral blood flow). Thus, this cross-sectional analysis was not specifically powered for the current analyses.

### Cardiopulmonary exercise test

CPET was performed following a standardized incremental ramp protocol on a treadmill, based on the American College of Sports Medicine guidelines.^36^ The exercise protocol consisted of: a 3-minute warm-up with speed progressively increasing until 4.8 km/h; the main part, consisting of 1% incline increments every 30 seconds until volitional exhaustion; and a 90-second active recovery. The maximal incline of the treadmill was 25%. The standardization of the protocol allowed using TTE as a maximal CPET indicator for comparison between subjects. Gas exchange was captured by a dilution flow system (Omnical, Maastricht Instruments, Maastricht, the Netherlands). A 12-lead electrocardiogram (ECG) (AMEDTEC ECGpro, GmbH, Aue, Germany) was continuously monitored by a cardiologist. Additionally, second-by-second heart rate (HR) was registered with a chest strap connected to a sports watch (Polar H10, Kempele, Finland). Every 3 minutes, blood pressure was measured with an automatic device (Tango® M2 ECG-gated Automated Blood Pressure Monitor, Suntech Medical Inc, NC, USA), and participants reported their rating of perceived exertion (RPE) on a visual OMNI scale (range 0-10).^37^ A more detailed description of the CPET protocol and data processing is published elsewhere.^38^

### CRF indicators

Peak heart rate (HR_peak_) was defined as the highest HR registered during the last 30 s of the CPET. Predicted HR_peak_ was calculated as 164-(0.7×age)^39^ for participants taking betablockers and as 211-(0.64×age) otherwise.^40^ Percentage of predicted HR_peak_ was then computed (HR_peak%pred_). Maximal indicators included VO_2peak_, TTE and peak oxygen pulse. VO_2peak_ (mL/kg/min) was defined as the highest 30-second average of absolute oxygen uptake during the CPET (mL/min) (this period could be extended to the active recovery period if needed) divided by body mass. Predicted VO_2peak_ was computed according to the equation from the FRIEND registry,^41^ and the percentage of predicted VO_2peak_ (VO_2peak%pred_) was calculated. Peak oxygen pulse was computed by dividing VO_2peak_ by HR_peak_. Submaximal indicators included HRrec, VAT and OUES. HRrec was calculated as the difference between HR_peak_ and HR after 60 s of active recovery. VAT (mL/kg/min of VO_2_) was determined with the V-slope method. OUES was obtained from the linear relationship between VO_2_ and log-transformed minute ventilation (VE), according to the following formula: VO_2_=*a* · log VE + *b*,^42^ where *a*=OUES and represents the slope of the model. Further description of the CRF indicators can be found in **Supplemental Table 1**.

### Life’s Essential 8 score

The LE8 score was calculated according to the AHA criteria.^6^ Scores from 0 to 100 were assigned to each individual component (see **Supplemental Table 2** for scoring details), and then the unweighted average was calculated to obtain the LE8 total score. The health behaviors score was computed as the average of sleep, nicotine exposure, physical activity and diet scores. The health factors score was computed as the average of BMI, blood lipids, blood glucose and blood pressure scores. Participants with valid data on <7 components were excluded.

### Health behaviors

Diet was measured with the Mediterranean Diet Adherence Screener (MEDAS) questionnaire (range 0-14 points).^43^ Since LE8 diet score is based on the Mediterranean Eating Pattern for Americans questionnaire^44^ (range 0-16 points),^6^ LE8 points were proportionally assigned to MEDAS points (see **Supplemental Table 3**). Self-reported nicotine exposure was collected retrospectively. Sleeping time and physical activity were assessed with an Axivity AX3 accelerometer (Axivity Ltd., New Castle Upon Tyne, United Kingdom) worn for 10 consecutive days. Raw accelerometer data were processed using the GGIR package^45^ (version 3.1.2, see https://github.com/Heart-Brain/Heart-Brain for the GGIR configuration file) in R (version 4.4.1), as previously described.^46^ Sleeping time was calculated as average hours of sleep per night. Physical activity was calculated as minutes of moderate-to-vigorous physical (i.e., accelerations above 100 m*g* (0.981m/s^2^)), only considering bouts longer than 1 minute.

### Health factors

Body mass and height were measured using a SECA 225 (Seca, Hamburg, Germany) stadiometer and scale. Fasting blood lipids, glucose and glycosylated hemoglobin (HbA1c) were determined from blood samples. Non-high-density lipoprotein (non-HDL) cholesterol was calculated by subtracting HDL cholesterol from total cholesterol. HbA1c measure was considered valid when assessed through the baseline blood draw or within the 90 previous days, in case the participant had no changes in glucose-related medication. Resting systolic and diastolic blood pressure were measured using an automated blood pressure monitor (Omron M3, Intellisense, OMRON Healthcare Europe, Spain) and the average of the three measures was computed for the analyses. Diabetes diagnoses and lipid-lowering and antihypertensive medications were gathered from participants’ medical records.

### Covariates

Sex assigned at birth, age at randomization time (calculated using birth date) and education level (primary, secondary or university studies) were self-reported by participants and used as covariates given their association with CRF and CVH metrics.^47–52^

### Statistical analysis

Data analyses and figures were conducted using R software (version 4.5.2). All statistical analyses were two-sided, considering *P*<.05 as statistically significant. Normality was tested using Shapiro-Wilk test. For sex, age groups and education groups comparison of continuous variables, Student’s t-test (equal variances) or Welch’s t-test (unequal variances) was used when data was normally distributed, and the Wilcoxon rank-sum test otherwise. Categorical variables were compared using the χ^2^ test of independence or the Fisher’s exact test.

Pearson correlations examined the unadjusted associations between covariates, CRF indicators and LE8 scores. Associations between CRF indicators and LE8 scores were examined using linear regression models adjusted for sex, age and education (primary, secondary or university) to address potential confounding. When data were missing, models included only participants with complete data for the variables analyzed. Sensitivity analyses were performed repeating the regression models in tests with a peak respiratory exchange ratio ≥1.10 and achieving ≥85% of HR_peak%pred_ to assess whether the associations for maximal indicators were altered. Linear regression results are presented as standardized beta (β_std_). The size of the associations was interpreted as small (0.10-0.29), medium (0.30-0.49) or large (≥0.50).^53^ Exploratory interaction analyses were conducted to evaluate whether associations between CPET indicators and LE8 total score differed by sex, age group (<65 years vs. ≥65 years), and education (primary/secondary vs. university). Sex-stratified analyses were adjusted for age and education, age-group-stratified analyses were adjusted for sex and education, and education stratified analyses were adjusted for sex and age.

## RESULTS

### Participant characteristics

Descriptive characteristics of the study sample are shown in **Table 1**. A total of 105 participants were enrolled in the Heart-Brain trial, and 102 (21 females, 62.3±6.6 years) were included in this study after excluding participants with incomplete LE8 data. Most of the participants had overweight (55.9%) or obesity (35.3%). Mean VO_2peak_ was 25.3±5.1 mL/kg/min, corresponding to 93.6±16.6% of the predicted value. The mean LE8 score was 66.6±8.6 points and there were no sex differences in the LE8 scores. LE8 individual component scores are displayed in **Figure 1**, ranging from 38.7±22.3 points for blood pressure to 94.1±16.4 points for physical activity. Age and education groups comparisons are presented in **Supplemental Tables 4 and 5**.

**Figure 1.**
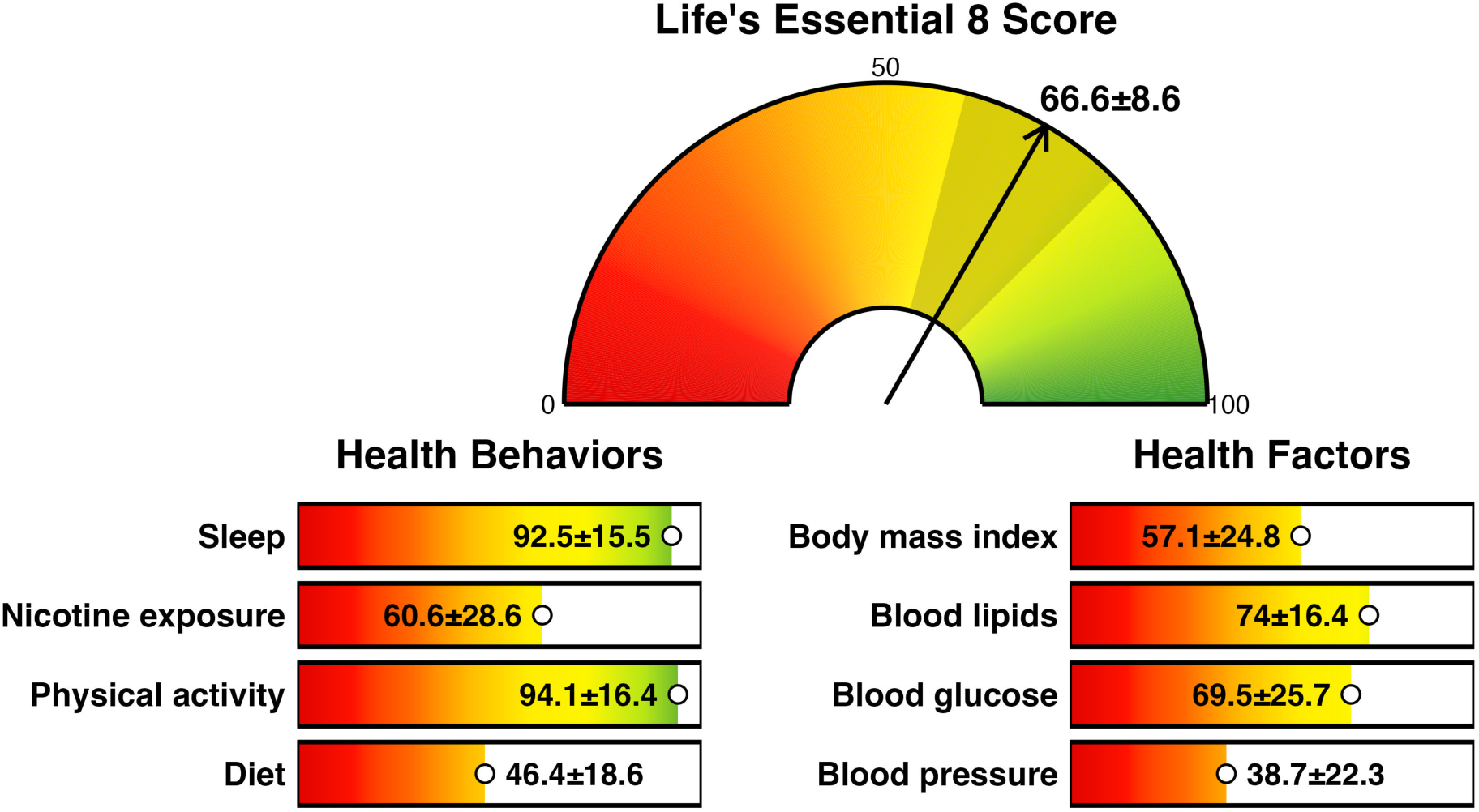
Scores (points) for overall cardiovascular health assessed by the Life’s Essential 8 and its individual components in the study sample. Data is presented as mean ± standard deviation. For the LE8 total score, the shaded area represents ± standard deviation.

**Table 1.** Descriptive characteristics of the study sample by sex.

|  | All<br>(n=102) | Males<br>(n=81) | Females<br>(n=21) | P-value |
| --- | --- | --- | --- | --- |
| **Demographics** |  |  |  |  |
| Age, years | 62.3 (6.6) | 62.4 (6.5) | 61.9 (7.1) | .77 |
| Body mass, kg | 83.1 (13.1) | 86.3 (11.5) | 70.7 (11.7) | <.001 |
| Height, cm | 168.9 (8.9) | 172.0 (6.6) | 156.7 (5.2) | <.001 |
| Education, categories |  |  |  | .21 |
| - Primary school | 20 (19.6%) | 13 (16.0%) | 7 (33.3%) |  |
| - Secondary school | 41 (40.2%) | 34 (42.0%) | 7 (33.3%) |  |
| - University | 41 (40.2%) | 34 (42.0%) | 7 (33.3%) |  |
| **Cardiovascular health-related factors** |  |  |  |  |
| Smoking, categories |  |  |  | <.001 |
| - Current smoker | 12 (11.9%) | 8 (9.9%) | 4 (20.0%) |  |
| - Former smoker | 75 (74.2%) | 67 (82.7%) | 8 (40.0%) |  |
| - Never smoker | 14 (13.9%) | 6 (7.4%) | 8 (40.0%) |  |
| MEDAS score (0-14 points) | 7.5 (1.9) | 7.2 (1.8) | 8.6 (2.0) | .006 |
| MVPA, min/week | 358.8 (273.5) | 381.1 (287.6) | 272.8 (192.3) | .09 |
| Sleep duration, h | 7.7 (1.0) | 7.7 (1.0) | 7.8 (1.2) | .97 |
| Body mass index, kg/m<sup>2</sup> | 29.1 (3.8) | 29.2 (3.7) | 28.8 (4.5) | .87 |
| Body mass index, categories |  |  |  | .005 |
| - Healthy weight | 9 (8.8%) | 3 (3.7%) | 6 (28.6%) |  |
| - Overweight | 57 (55.9%) | 50 (61.7%) | 7 (33.3%) |  |
| - Obesity class I | 27 (26.5%) | 21 (25.9%) | 6 (28.6%) |  |
| - Obesity class II | 9 (8.8%) | 7 (8.6%) | 2 (9.5%) |  |
| Diabetes mellitus type II | 18 (17.6%) | 16 (19.8%) | 2 (9.5%) | .35 |
| Fasting blood glucose, mg/dL | 102.9 (29.1) | 106.0 (30.0) | 91.0 (21.8) | <.001 |
| HbA1c, % | 5.8 (0.8) | 5.9 (0.8) | 5.5 (0.4) | .35 |
| Non-HDL cholesterol, mg/dL | 88.4 (32.8) | 83.9 (28.5) | 105.8 (41.9) | .04 |
| Systolic BP, mmHg | 126.9 (14.5) | 126.9 (13.3) | 126.8 (18.8) | .97 |
| Diastolic BP, mmHg | 78.3 (9.0) | 78.0 (8.4) | 79.7 (11.1) | .43 |
| Anti-hypertensive medication | 83 (81.4%) | 67 (82.7%) | 16 (76.2%) | .71 |
| Lipid-lowering medication | 101 (99.0%) | 81 (100%) | 20 (95.2%) | .21 |
| Betablockers | 77 (75.5%) | 63 (77.8%) | 14 (66.7%) | .44 |
| **LE8 score** (0-100 points) | 66.6 (8.6) | 66.4 (8.6) | 67.4 (9.1) | .63 |
| **Health behaviors score**, range 0-100 points | 73.5 (10.3) | 72.9 (9.7) | 75.9 (12.4) | .15 |
| **Health factors score**, range 0-100 points | 59.6 (14.1) | 59.8 (13.8) | 58.9 (15.4) | .81 |
| **Cardiopulmonary exercise test** |  |  |  |  |
| VO<sub>2peak</sub>, mL/kg/min | 25.3 (5.1) | 26.3 (4.9) | 21.7 (4.1) | <.001 |
| VO<sub>2peak</sub>, mL/kg/min, % pred | 93.6 (16.6) | 88.8 (11.9) | 111.8 (19.2) | <.001 |
| Time to exhaustion, s | 661.8 (128.7) | 684.7 (122.1) | 573.4 (116.8) | <.001 |
| HR<sub>peak</sub>, beats/min | 138.2 (17.0) | 138.3 (17.3) | 137.8 (15.9) | .71 |
| HR<sub>peak</sub>, % pred HR<sub>peak</sub> | 105.7 (15.5) | 106.7 (15.9) | 102.1 (13.6) | .24 |
| HR<sub>peak</sub>, ≥ 85% pred HR<sub>peak</sub> | 94 (92.2%) | 75 (92.6%) | 19 (90.5%) | .67 |
| RER<sub>peak</sub> | 1.183 (0.1) | 1.195 (0.1) | 1.139 (0.1) | .02 |
| RER<sub>peak</sub> ≥ 1.10, n (%) | 77 (75.5%) | 64 (79.0%) | 13 (61.9%) | .08 |
| RPE<sub>peak</sub>, range 0-10 | 7.7 (1.3) | 7.6 (1.3) | 7.9 (1.5) | .61 |
| HR<sub>rec</sub>, beats/min | 21.6 (8.1) | 21.2 (7.9) | 23.2 (9.0) | .34 |
| VAT, mL/kg/min of VO<sub>2</sub> | 16.8 (3.4) | 17.3 (3.3) | 14.9 (2.9) | .002 |
| OUES, mL/min/log(L/min) | 765.6 (222.7) | 824.8 (189.6) | 537.3 (194.1) | <.001 |
| VO<sub>2</sub> pulse, mL/beat | 15.2 (3.1) | 16.3 (2.4) | 11.0 (1.6) | <.001 |
| Systolic BP<sub>peak</sub>, mmHg | 183.9 (23.8) | 183.5 (21.9) | 185.7 (31.1) | .77 |

Data is presented as mean (SD) for continuous variables and as n (%) for categorical variables. Due to missing data, smoking status: n=101 (males= 81, females=20), non-HDL cholesterol: n=96 (males=76, females=20), VO<sub>2peak</sub>: n=100 (males=79, females=21), RER<sub>peak</sub>: n=95 (males=75, females=20), RPE<sub>peak</sub>: n=98 (males=81, females=17), HR<sub>rec</sub>: n=99 (males=80, females=19), VAT: n=96 (males=76, females=20), VO<sub>2</sub> pulse: n=100 (males=79, females=21), SBP<sub>peak</sub>: n=98 (males=79, females=19).
Abbreviations: %pred, percent of predicted; BP, blood pressure; HbA1c, glycosylated hemoglobin; HR, heart rate; HR<sub>rec</sub>, heart rate recovery; LE8, Life's Essential 8; MEDAS, Mediterranean Diet Adherence Screener; MVPA, moderate-to-vigorous intensity physical activity; non-HDL, non-high-density lipoprotein; OUES, oxygen uptake efficiency slope; RER, respiratory exchange ratio; RPE, rating of perceived exertion; VAT, ventilatory anaerobic threshold; VO<sub>2</sub>, oxygen uptake.

### Correlation analyses

**Figure 2** presents Pearson correlation results between covariates, CRF indicators and LE8 scores. TTE had small to medium size correlations with LE8, health behaviors and health factors scores (r=0.25 to 0.43, all *P*<.01), while VO_2peak_, VAT and HRrec showed correlation with LE8 and health factors (r=0.24 to 0.47, all *P*<.02). No significant correlations were found between peak oxygen pulse and OUES with LE8 scores.

**Figure 2.**
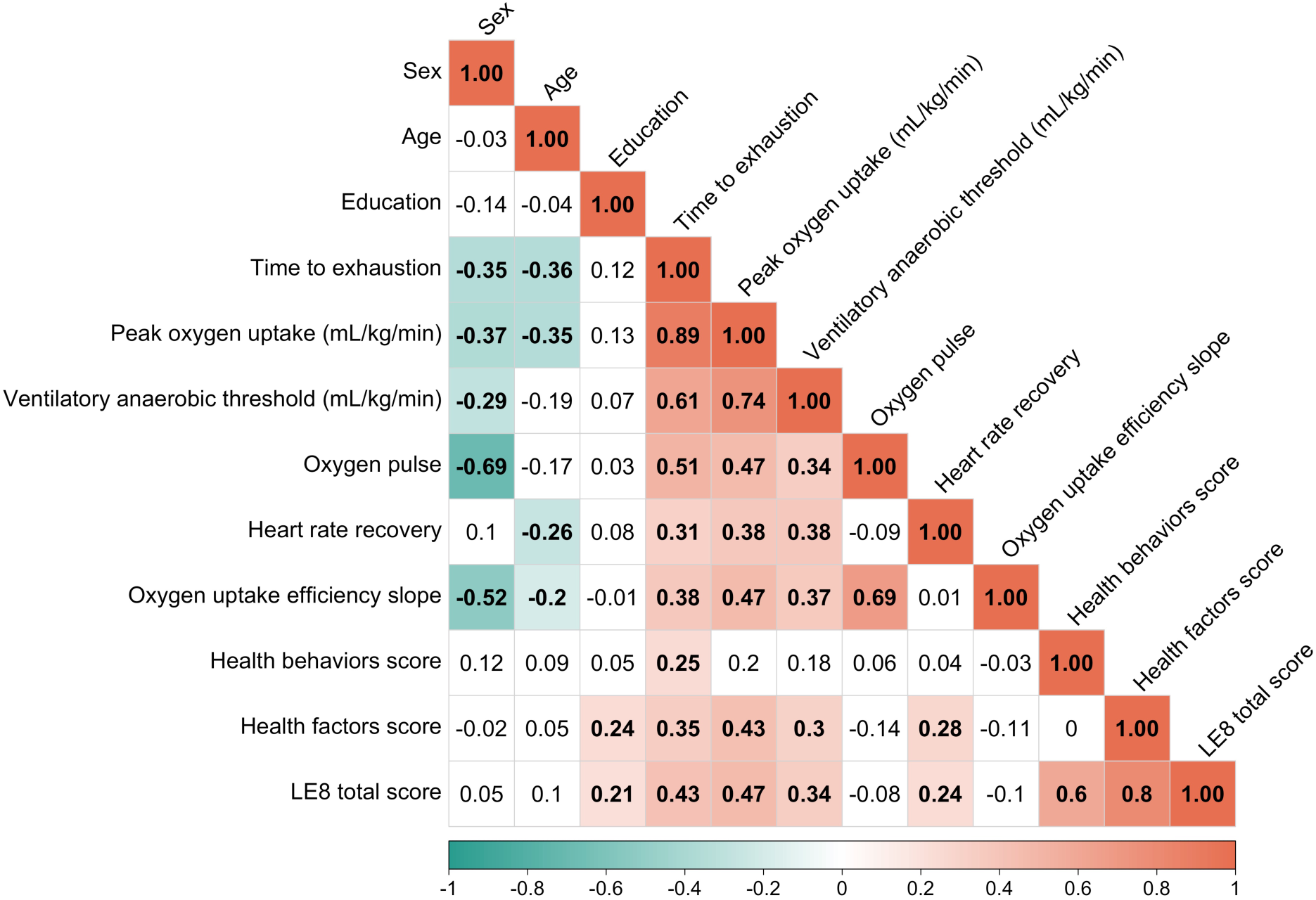
Pearson unadjusted correlation matrix (coefficients shown, r) between cardiopulmonary exercise test indicators, Life’s Essential 8 scores and covariates. Statistically significant correlations (*P*<.05) are presented in bold and colored. The colors of the boxes represent the strength and direction of the significant correlations. Sex was coded as 0 = male and 1 = female. Education was coded as 1 = primary studies, 2 = secondary studies, and 3 = university studies. Exact *P* values for correlation analyses can be found in **Supplemental Table 6**.

### Linear regression analyses

Results of linear regression models adjusted by sex, age and education are shown in **Figure 3**. Unstandardized coefficients are shown in **Supplemental Table 7**. VO_2peak_ (β_std_=0.67, *P*<.001) and TTE (β_std_=0.64, *P*<.001) showed the strongest positive associations with LE8 total score, followed by VAT (β_std_=0.43, *P*<.001) and HRrec (β_std_=0.26, *P*=.01). No statistically significant associations were observed for OUES (β_std_=-0.08, *P*=.54) and peak oxygen pulse (β_std_=-0.05, *P*=.72) with LE8 total score. VO_2peak_, TTE and VAT were also significantly associated with both health behaviors and health factors (β_std_=0.27-0.56, all *P*<.01), whereas HRrec was only significantly associated with health factors (β_std_=0.30, *P*=.004). Peak oxygen pulse was positively associated with health behaviors (β_std_=0.32, *P*=.03) and negatively associated with health factors (β_std_=-0.31, *P*=.03). The associations between maximal fitness indicators and LE8 total score were overall consistent when the analysis was restricted to those CPETs in which respiratory exchange ratio was ≥1.10 and HR_peak%pred_ was ≥85% (n=76) (see **Supplemental Table 8**).

**Figure 3.**
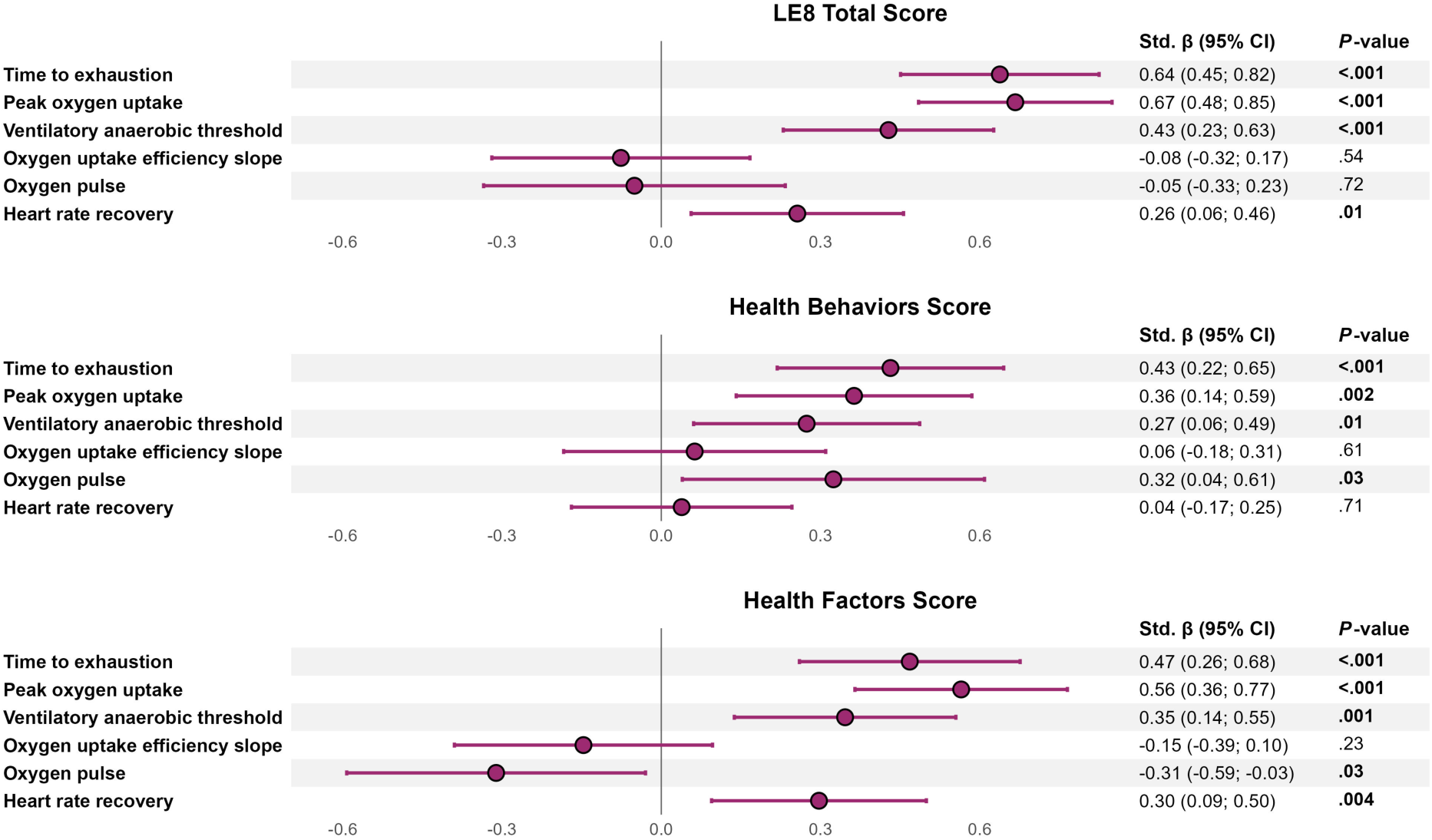
Linear regressions between cardiorespiratory fitness indicators and Life’s Essential 8 scores. The results represent standardized beta coefficients from linear regression analyses, with 95% confidence intervals and *P*-values. All analyses were adjusted for sex, age and education. Significant *P*-values are presented in bold. Abbreviations: CI, confidence interval; LE8, Life’s Essential 8.

### Exploratory sub-group analyses

**Figure 4** shows exploratory analyses to examine the interaction by sex, age group, and education in the associations between CRF indicators and LE8 total score. VO_2peak_ and TTE consistently showed positive, large significant associations with LE8 across sex, age and education groups (β_std_=0.51-0.92, all *P*<.005). No statistically significant interaction was observed by sex or education. For age groups, a trend towards stronger associations in older participants was observed for most of CRF indicators, yet significant interactions were observed only for VAT (*P*-int=.03) and OUES (*P*-int=.02).

**Figure 4.**
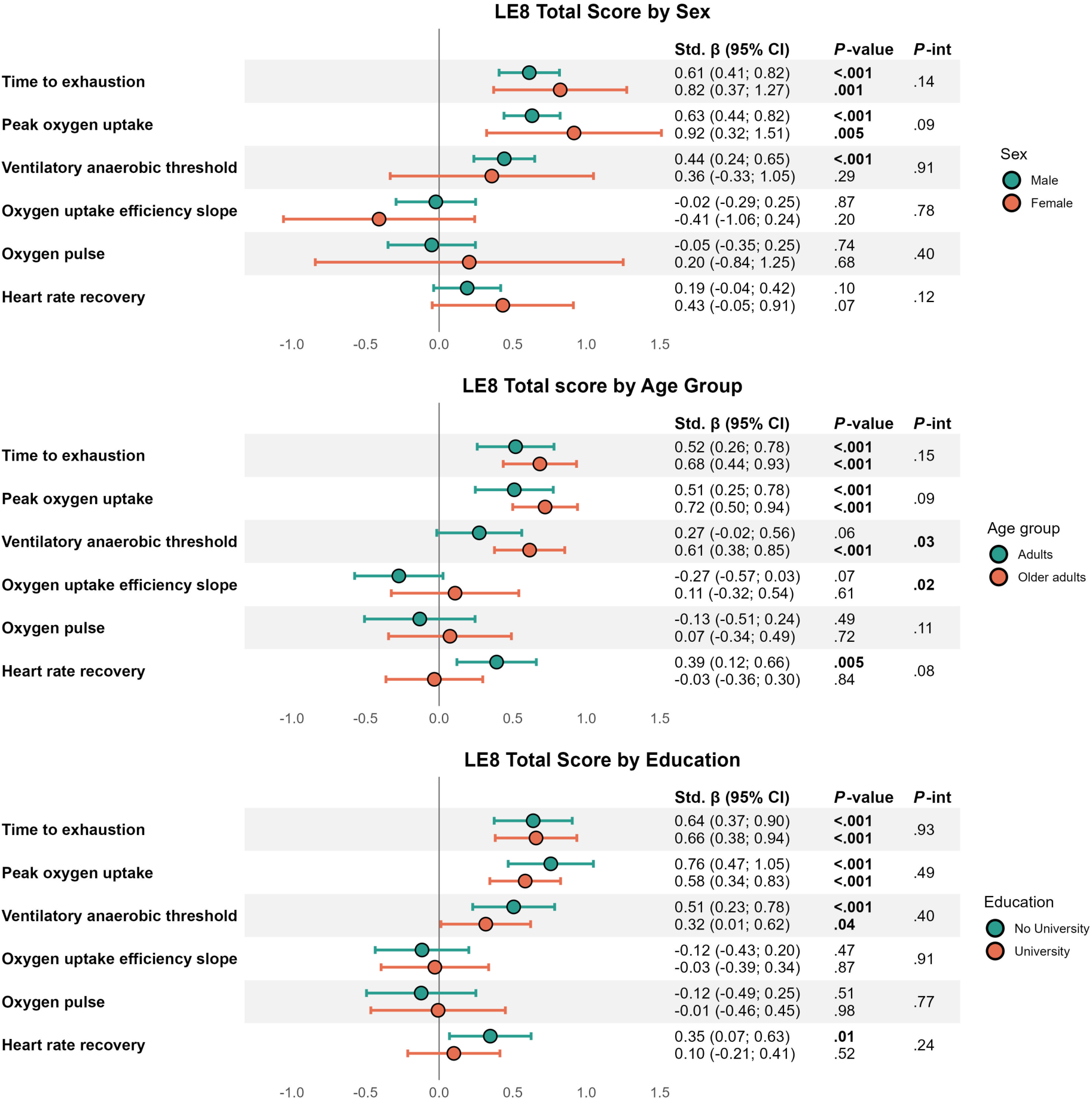
Linear regressions between cardiorespiratory fitness indicators and LE8 total score, stratified by sex, age group and educational level. The results represent standardized beta coefficients from linear regression analyses, with 95% confidence intervals. Estimates in the sex-stratified analysis are adjusted for age and education; estimates in the age-group–stratified (<65 years old vs ≥65 years old) analysis are adjusted for sex and education; and estimates in the education-stratified analysis are adjusted for sex and age. *P*-int indicates the *P*-value for interaction between groups. Significant *P*-values are represented in bold. Abbreviations: CI, confidence interval; LE8, Life’s Essential 8.

## DISCUSSION

Our findings support that: (i) TTE, VO_2peak_, VAT and HRrec are significantly and positively associated with LE8 total score after adjusting for potential confounders (i.e. sex, age and education) in individuals with CAD; (ii) the effect sizes of the associations were markedly larger for maximal CRF indicators (TTE and VO_2peak_) than for submaximal indicators, with medium effect size for VAT, and small for HRrec; (iii) TTE notably demonstrated a similar strength of association with LE8 as VO_2_peak, the gold-standard measure of CRF, highlighting the potential value of TTE as a simpler (given its greater feasibility for routine clinical implementation) yet informative marker of CVH in individuals with CAD; and (iv) the associations for VO_2peak_ and TTE were consistently significant in males and females, younger and older participants, and across educational levels.

These findings are in line with a substantial body of evidence that positions CRF as a key marker of CVH.^16^ However, the relationship between CRF and CVH as assessed with LE8 has been scarcely investigated.^34,54^ This focus on LE8 is particularly relevant because, compared with the Life’s Simple 7, the previous CVH construct, LE8 has shown slightly better discriminatory capacity for coronary stenosis,^7^ suggesting that it may capture CVH with greater clinical relevance in populations with CAD, in whom stenotic coronary disease is highly prevalent. Moreover, to the best of our knowledge, no previous study has specifically examined these associations in individuals with stable CAD. By examining multiple CRF indicators, our study extends prior research to a secondary prevention setting and provides new insights into how different aspects of CRF relate to CVH in individuals with CAD.

In our study, maximal CRF indicators (VO_2peak_ and TTE) showed the largest positive associations with LE8. The scarcity of CAD-specific studies precludes direct comparison with prior studies in this population, but our findings suggest that the relationship between maximal indicators of CRF and LE8 in individuals with CAD is consistent with prior work in the general population. Ravichandran et al. reported that a clinically meaningful 5-point higher LE8 score was associated with a 6% greater VO_2peak_ in a community-based sample.^34^ Notably, they also observed that this association was consistent among participants with prevalent cardiovascular disease (n=68). In the Aerobics Center Longitudinal Study (n=11,590 adults), CRF estimated by maximal treadmill time was positively correlated with Life’s Simple 7 in males (r=0.56) and females (r=0.50), similar to our findings between TTE and LE8 (r=0.43). Longitudinal studies also support a positive relationship between maximal exercise capacity and Life’s Simple 7/LE8 in different populations.^33,34,54^ Because maximal exercise capacity integrates central (cardiovascular and pulmonary) and peripheral (skeletal muscle) function and has consistently shown prognostic value across diverse cardiovascular and pulmonary conditions,^55^ its close relationship with LE8 likely reflects overall disease burden in individuals with CAD. Furthermore, the similar strength of association observed for TTE and VO_2peak_ is clinically relevant. While VO_2peak_ remains the gold-standard measure of CRF, it requires gas-exchange analysis, specialized equipment, and greater data processing. In contrast, TTE can be obtained from a standardized maximal exercise test without gas-exchange, which may make it a simpler marker for monitoring CVH in clinical settings.

Beyond maximal indicators, VAT and HRrec were also positively associated with LE8 total score. Although, to our knowledge, there is no prior evidence linking VAT and HRrec to LE8, both measures may reflect CVH since they have been inversely associated with mortality risk,^56–59^ even in populations with cardiovascular disease.^59,60^ This could have clinical implications, since VAT can be identified without requiring volitional exhaustion, and HRrec is particularly feasible for clinical use because it only requires HR monitoring during recovery. Peak oxygen pulse was not associated with LE8 total score, but it showed positive associations with health behaviors and negative associations with health factors. Prior evidence linking peak oxygen pulse to higher physical activity level as an indicator of maximal left ventricular stroke volume during exercise and to lower nicotine exposure levels may explain the connection with health behaviors.^61–63^ The inverse relationship with health factors may partly be explained by body size, as O_2_ pulse is derived from absolute VO_2peak_ (mL/min). Larger individuals typically have higher absolute VO_2peak_ and, consequently, higher peak oxygen pulse, while greater adiposity may contribute to lower LE8 health factors scores through BMI and potentially less favorable cardiometabolic profile. OUES showed no association with LE8 scores in our cohort, in contrast with previous evidence supporting OUES as an independent prognostic marker in CAD.^23,27^

In exploratory subgroup analyses, associations between CRF indicators and LE8 were generally consistent across sex, age groups, and educational levels. A trend towards stronger associations in older participants was observed for most of CRF indicators, with significant interactions observed only for Age x VAT and Age x OUES. These significant interactions suggest potential age-related heterogeneity, although these findings should be confirmed in larger sample sizes, given our limited subgroup sample sizes and multiple testing performed. The most consistent associations were observed for TTE and VO_2peak_, which were positively associated with LE8 across all subgroups, reinforcing the consistency of the findings for these two indicators.

### Strengths and limitations

Strengths of this study include: the comprehensive assessment of CRF measures obtained from a standardized CPET, allowing a broad characterization of cardiovascular and respiratory functional status; the objective assessment of physical activity and sleep using accelerometry, reducing potential reporting bias; and the use of LE8 as a comprehensive, continuous measure of CVH that integrates both health behaviors and cardiometabolic health factors. However, some limitations should be acknowledged. The cross-sectional design precludes causal inference and the multiple tests performed may increase the likelihood of type I errors. Subgroup analyses should be interpreted cautiously due to the limited sample size, particularly among females (n=21). Finally, as data came from a trial designed to safely deliver high-intensity interval training, participants were likely healthier and more functionally preserved than the average CAD population, limiting the generalizability to less active or more severely affected individuals with CAD.

## CONCLUSION

In individuals with CAD, higher maximal and submaximal CRF indicators were associated with a higher LE8 total score. Associations were the strongest for the maximal indicators TTE and VO_2peak_, while VAT and HRrec showed weaker, yet significant, associations. Notably, TTE demonstrated an association with LE8 of comparable magnitude to that of VO_2peak_, which has important clinical implications, underscoring its value as a simple and powerful health indicator that requires less equipment, examiner training and data processing. Importantly, these associations were significant in females and males, younger and older and individuals with higher and lower educational level, underscoring the robustness of the findings.

## Data Availability

Data are available upon reasonable request to the authors

https://github.com/Heart-Brain/Heart-Brain

## ACKNOWLEDGEMENTS

This work is part of a Ph.D. thesis conducted in the Biomedicine Doctoral Studies of the University of Granada, Spain. We would like to thank all participants for their time, effort and valuable contribution to this study.

## ETHICS STATEMENT

This study used baseline data from the Heart-Brain trial (NCT06214624) and was approved by the Research Ethics Board of the Andalusian Health Service (CEIM/CEI Provincial de Granada; #1776-N-21). All participants provided written informed consent.

## ABBREVIATIONS

AHA: American Heart Association
BMI: body mass index
CAD: coronary artery disease
CPET: cardiopulmonary exercise test
CRF: cardiorespiratory fitness
CVH: cardiovascular health
HDL: high-density lipoprotein
HR: heart rate
HRrec: heart rate recovery
LE8: Life’s Essential 8
MEDAS: mediterranean diet adherence screener
OUES: oxygen uptake efficiency slope
TTE: time to exhaustion
VAT: ventilatory anaerobic threshold
VO_2peak_: peak oxygen uptake

## SUPPLEMENTAL MATERIAL

STROBE Statement. Checklist of items that should be included in reports of observational studies

|  | **Item**<br>**No.** | **Recommendation** | **Page**<br>**No.** |
| --- | --- | --- | --- |
| **Title and abstract** | 1 | (a) Indicate the study's design with a commonly used term in the title or the abstract | 4 |
|  |  | (b) Provide in the abstract an informative and balanced summary of what was done and what was found | 4 |
| **Introduction** |  |  |  |
| Background/rationale | 2 | Explain the scientific background and rationale for the investigation being reported | 7, 8 |
| Objectives | 3 | State specific objectives, including any prespecified hypotheses | 8 |
| **Methods** |  |  |  |
| Study design | 4 | Present key elements of study design early in the paper | 8, 9 |
| Setting | 5 | Describe the setting, locations, and relevant dates, including periods of recruitment, exposure, follow-up, and data collection | 9 |
| Participants | 6 | (a) Give the eligibility criteria, and the sources and methods of selection of participants | 9, Ref 35 |
| Variables | 7 | Clearly define all outcomes, exposures, predictors, potential confounders, and effect modifiers. Give diagnostic criteria, if applicable | 10-13 |
| Data sources/<br>measurement | 8 | For each variable of interest, give sources of data and details of methods of assessment (measurement). Describe comparability of assessment methods if there is more than one group | 10-12 |
| Bias | 9 | Describe any efforts to address potential sources of bias | 12 |
| Study size | 10 | Explain how the study size was arrived at | 9, Ref 35 |
| Quantitative<br>variables | 11 | Explain how quantitative variables were handled in the analyses. If applicable, describe which groupings were chosen and why | 12-13 |
| Statistical methods | 12 | (a) Describe all statistical methods, including those used to control for confounding | 12-13 |
|  |  | (b) Describe any methods used to examine subgroups and interactions | 12-13 |
|  |  | (c) Explain how missing data were addressed | 12 |
|  |  | (d) If applicable, describe analytical methods taking account of sampling strategy | NA |
|  |  | (e) Describe any sensitivity analyses | 12 |
| Results |  |  |  |
| Participants | 13 | (a) Report numbers of individuals at each stage of study—eg numbers potentially eligible, examined for eligibility, confirmed eligible, included in the study, completing follow-up, and analysed | 13 |
|  |  | (b) Give reasons for non-participation at each stage | 13 |
|  |  | (c) Consider use of a flow diagram | NA |
| Descriptive data | 14 | (a) Give characteristics of study participants (eg demographic, clinical, social) and information on exposures and potential confounders | 13 |
|  |  | (b) Indicate number of participants with missing data for each variable of interest | 13, Table 1, Supplemental Tables 5 and 6 |
| Outcome data | 15 | Report numbers of outcome events or summary measures | 13, Table 1 |
| Main results | 16 | (a) Give unadjusted estimates and, if applicable, confounder-adjusted estimates and their precision (eg, 95% confidence interval). Make clear which confounders were adjusted for and why they were included | Figures 2 and 3 |
|  |  | (b) Report category boundaries when continuous variables were categorized | 13 |
|  |  | (c) If relevant, consider translating estimates of relative risk into absolute risk for a meaningful time period | NA |
| Other analyses | 17 | Report other analyses done—eg analyses of subgroups and interactions, and sensitivity analyses | 14 |
| **Discussion** |  |  |  |
| Key results | 18 | Summarise key results with reference to study objectives | 14, 15 |
| Limitations | 19 | Discuss limitations of the study, taking into account sources of potential bias or imprecision. Discuss both direction and magnitude of any potential bias | 17, 18 |
| Interpretation | 20 | Give a cautious overall interpretation of results considering objectives, limitations, multiplicity of analyses, results from similar studies, and other relevant evidence | 15-17 |
| Generalisability | 21 | Discuss the generalisability (external validity) of the study results | 17, 18 |
| **Other information** |  |  |  |
| Funding | 22 | Give the source of funding and the role of the funders for the present study and, if applicable, for the original study on which the present article is based | 2, 3 |

**Supplemental Table 1.**
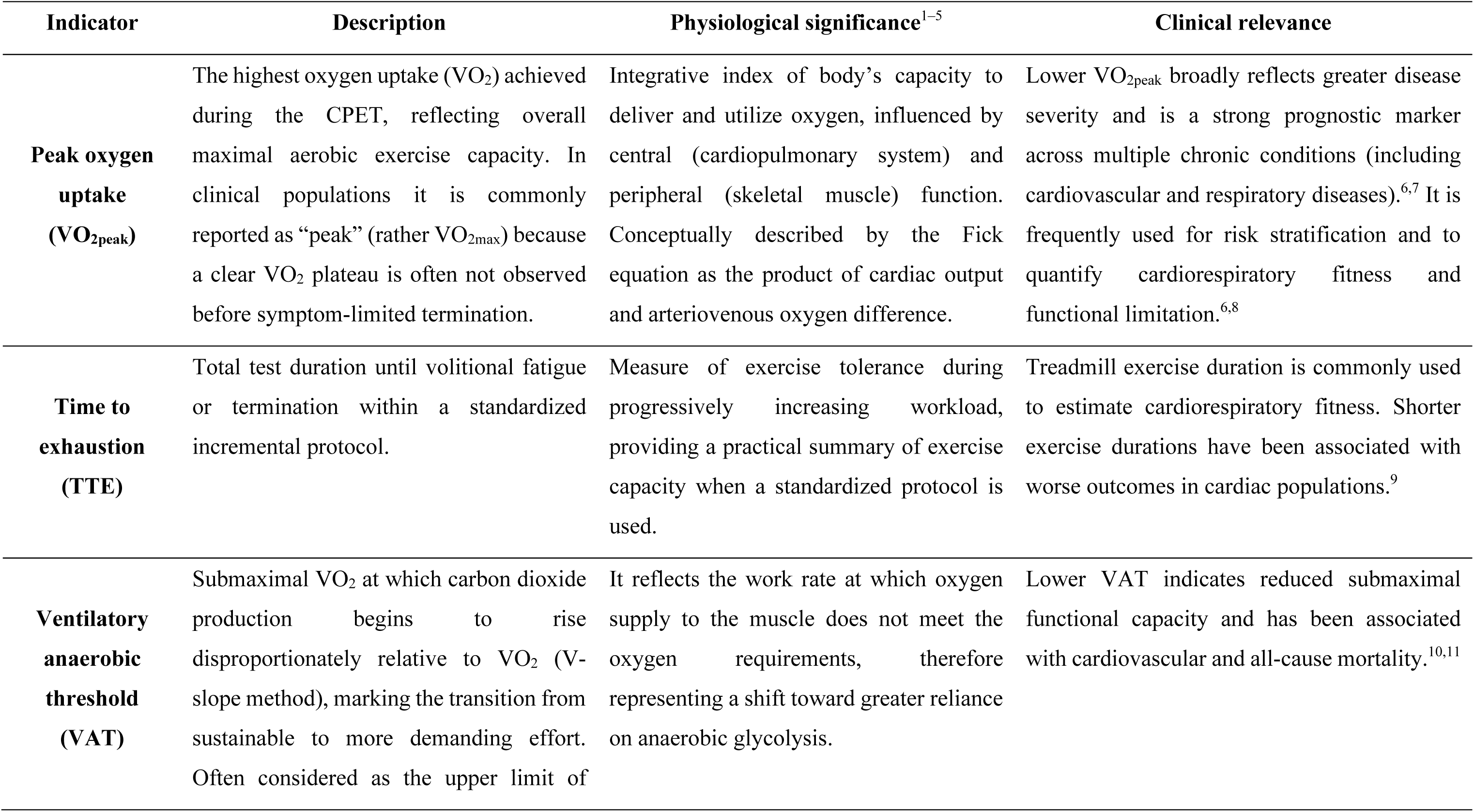
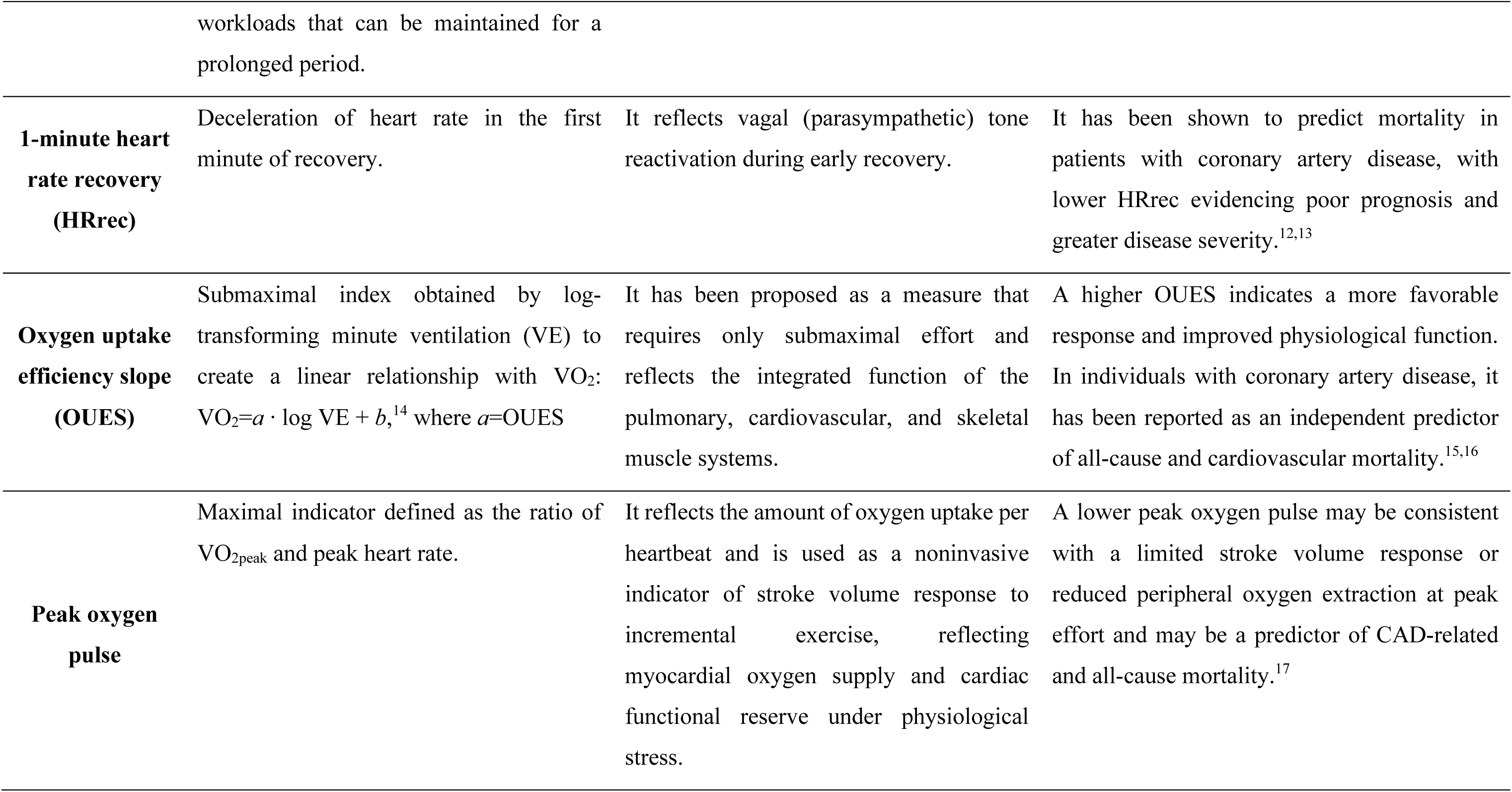
Cardiorespiratory fitness (CRF) indicators obtained from the cardiopulmonary exercise test (CPET).

**Supplemental Table 2.** Life’s Essential 8 calculation.

| CVH metric | Quantification of CVH metric | LE8 Points | All (n=102) | Male (n=81) | Female (n=21) | Adults (n=65) | Older adults (n=37) |
| --- | --- | --- | --- | --- | --- | --- | --- |
| Diet: Mediterranean Diet Adherence Screener questionnaire (MEDAS) | 0 – 2 MEDAS points | 0 | 0 (0%) | 0 (0%) | 0 (0%) | 0 (0%) | 0 (0%) |
|  | 3 – 6 MEDAS points | 25 | 33 (32.4%) | 32 (39.5%) | 1 (4.8%) | 26 (40.0%) | 7 (18.9%) |
|  | 7 – 9 MEDAS points | 50 | 55 (53.9%) | 41 (50.6%) | 14 (66.7%) | 33 (50.8%) | 22 (59.5%) |
|  | 10 – 12 MEDAS points | 80 | 12 (11.8%) | 7 (8.6%) | 5 (23.8%) | 5 (7.7%) | 7 (18.9%) |
|  | 13 – 14 MEDAS points | 100 | 2 (2.0%) | 1 (1.2%) | 1 (4.8%) | 1 (1.5%) | 1 (2.7%) |
| Physical activity: Moderate to vigorous physical activity (MVPA) minutes assessed with accelerometry, with 1-minute frequency | 0 min of MVPA per week | 0 | 0 (0%) | 0 (0%) | 0 (0%) | 0 (0%) | 0 (0%) |
|  | 1–29 min of MVPA per week | 20 | 2 (2.0%) | 0 (0%) | 2 (9.5%) | 0 (0%) | 2 (5.4%) |
|  | 30–59 min of MVPA per week | 40 | 3 (2.9%) | 3 (3.7%) | 0 (0%) | 2 (3.1%) | 1 (2.7%) |
|  | 60–89 min of MVPA per week | 60 | 3 (2.9%) | 2 (2.5%) | 1 (4.8%) | 0 (0%) | 3 (8.1%) |
|  | 90–119 min of MVPA per week | 80 | 5 (4.9%) | 4 (4.9%) | 1 (4.8%) | 4 (6.2%) | 1 (2.7%) |
|  | 120–149 min of MVPA per week | 90 | 4 (3.9%) | 1 (1.2%) | 3 (14.3%) | 1 (1.5%) | 3 (8.1%) |
|  | ≥150 min of MVPA per week | 100 | 85 (83.3%) | 71 (87.7%) | 14 (66.7%) | 58 (89.2%) | 27 (73.0%) |
| Nicotine exposure: Self-reported use of cigarettes or inhaled nicotine delivery systems | Current smoker | 0 | 12 (11.8%) | 8 (9.9%) | 4 (19.0%) | 9 (13.8%) | 3 (8.1%) |
|  | Former smoker, quit <1 y, living with indoor smoker | 5 | 1 (1.0%) | 1 (1.2%) | 0 (0%) | 1 (1.5%) | 0 (0%) |
|  | Former smoker, quit <1 y, or using inhaled NDS | 25 | 0 (0%) | 0 (0%) | 0 (0%) | 0 (0%) | 0 (0%) |
|  | Former smoker, quit 1-<5 y, living with indoor smoker | 30 | 4 (3.9%) | 4 (4.9%) | 0 (0%) | 4 (6.2%) | 0 (0%) |
|  | Former smoker, quit 1-<5 y | 50 | 19 (18.6%) | 16 (19.8%) | 3 (14.3%) | 13 (20.0%) | 6 (16.2%) |
|  | Former smoker, quit ≥5 y, living with indoor smoker | 55 | 7 (6.9%) | 7 (8.6%) | 0 (0%) | 4 (6.2%) | 3 (8.1%) |
|  | Former smoker, quit ≥5 y | 75 | 44 (43.1%) | 39 (48.1%) | 5 (23.8%) | 25 (38.5%) | 19 (51.4%) |
|  | Never smoker, living with indoor smoker | 80 | 2 (2.0%) | 1 (1.2%) | 1 (4.8%) | 2 (3.1%) | 0 (0%) |
|  | Never smoker | 100 | 12 (11.8%) | 5 (6.2%) | 7 (33.3%) | 6 (9.2%) | 6 (16.2%) |
| Sleep:<br>Hours of sleep per night assessed with accelerometry | <4 h | 0 | 0 (0%) | 0 (0%) | 0 (0%) | 0 (0%) | 0 (0%) |
|  | 4-<5 h | 20 | 0 (0%) | 0 (0%) | 0 (0%) | 0 (0%) | 0 (0%) |
|  | 5-<6 or >10 h | 40 | 5 (4.9%) | 3 (3.7%) | 2 (9.5%) | 3 (4.6%) | 2 (5.4%) |
|  | 6-<7 h | 70 | 12 (11.8%) | 10 (12.3%) | 2 (9.5%) | 8 (12.3%) | 4 (10.8%) |
|  | 9-<10 h | 90 | 10 (9.8%) | 9 (11.1%) | 1 (4.8%) | 3 (4.6%) | 7 (18.9%) |
|  | 7-<9 h | 100 | 75 (73.5%) | 59 (72.8%) | 16 (76.2%) | 51 (78.5%) | 24 (64.9%) |
| BMI:<br>Body mass (kilograms) divided by height squared (meters squared). Measured by research staff | Obesity III | 0 | 0 (0%) | 0 (0%) | 0 (0%) | 0 (0%) | 0 (0%) |
|  | Obesity II | 15 | 9 (8.8%) | 7 (8.6%) | 2 (9.5%) | 7 (10.8%) | 2 (5.4%) |
|  | Obesity I | 30 | 27 (26.5%) | 21 (25.9%) | 6 (28.6%) | 19 (29.2%) | 8 (21.6%) |
|  | Overweight | 70 | 57 (55.9%) | 50 (61.7%) | 7 (33.3%) | 35 (53.8%) | 22 (59.5%) |
|  | Healthy weight | 100 | 9 (8.8%) | 3 (3.7%) | 6 (28.6%) | 4 (6.2%) | 5 (13.5%) |
| Blood lipids:<br>non-HDL cholesterol calculated based on fasting plasma total cholesterol and HDL cholesterol | >220 or (190–219 mg/dL and taking medication) | 0 | 0 (0%) | 0 (0%) | 0 (0%) | 0 (0%) | 0 (0%) |
|  | 190–219 mg/dL (or 160–189 mg/dL and taking medication) | 20 | 5 (4.9%) | 1 (1.2%) | 4 (19.0%) | 2 (3.1%) | 3 (8.1%) |
|  | 160–189 or (130–159 mg/dL and taking medication) | 40 | 7 (6.9%) | 5 (6.2%) | 2 (9.5%) | 7 (10.8%) | 0 (0%) |
|  | 130–159 mg/dL without lipid-lowering medication | 60 | 0 (0%) | 0 (0%) | 0 (0%) | 0 (0%) | 0 (0%) |
|  | <130 mg/dL and taking medication | 80 | 84 (82.4%) | 70 (86.4%) | 14 (66.7%) | 54 (83.1%) | 30 (81.1%) |
|  | <130 mg/dL without lipid-lowering medication | 100 | 0 (0%) | 0 (0%) | 0 (0%) | 0 (0%) | 0 (0%) |
| Blood glucose:<br>Fasting blood glucose (FBG, mg/dL) or HbA1c (%) | Diabetes with HbA1c ≥10.0 | 0 | 0 (0%) | 0 (0%) | 0 (0%) | 0 (0%) | 0 (0%) |
|  | Diabetes with HbA1c 9.0–9.9 | 10 | 2 (2.0%) | 2 (2.5%) | 0 (0%) | 1 (1.5%) | 1 (2.7%) |
|  | Diabetes with HbA1c 8.0–8.9 | 20 | 2 (2.0%) | 1 (1.2%) | 1 (4.8%) | 1 (1.5%) | 1 (2.7%) |
|  | Diabetes with HbA1c 7.0–7.9 | 30 | 4 (3.9%) | 4 (4.9%) | 0 (0%) | 3 (4.6%) | 1 (2.7%) |
|  | Diabetes with HbA1c <7.0 | 40 | 11 (10.8%) | 9 (11.1%) | 2 (9.5%) | 9 (13.8%) | 2 (5.4%) |
|  | No diabetes and FBG 100–125 (or HbA1c 5.7–6.4) | 60 | 45 (44.1%) | 36 (44.4%) | 9 (42.9%) | 28 (43.1%) | 17 (45.9%) |
|  | No diabetes and FBG <100 (or HbA1c <5.7) | 100 | 37 (36.3%) | 28 (34.6%) | 9 (42.9%) | 23 (35.4%) | 14 (37.8%) |
| Blood pressure (BP):<br>Resting systolic and diastolic BPs (mmHg) measured with an automated BP monitor | ≥160 or ≥100 | 0 | 3 (2.9%) | 1 (1.2%) | 2 (9.5%) | 2 (3.1%) | 1 (2.7%) |
|  | (140–150 or 90–99) and taking medication | 5 | 19 (18.6%) | 15 (18.5%) | 4 (19.0%) | 13 (20.0%) | 6 (16.2%) |
|  | 140–150 or 90–99 | 25 | 3 (2.9%) | 2 (2.5%) | 1 (4.8%) | 2 (3.1%) | 1 (2.7%) |
|  | (130–139 or 80–89) and taking medication | 30 | 23 (22.5%) | 19 (23.5%) | 4 (19.0%) | 14 (21.5%) | 9 (24.3%) |
|  | 130–139 or 80–89 | 50 | 8 (7.8%) | 7 (8.6%) | 1 (4.8%) | 3 (4.6%) | 5 (13.5%) |
|  | 120–129/<90 and taking medication | 55 | 38 (37.3%) | 32 (39.5%) | 6 (28.6%) | 25 (38.5%) | 13 (35.1%) |
|  | 120–129/<90 | 75 | 8 (7.8%) | 5 (6.2%) | 3 (14.3%) | 6 (9.2%) | 2 (5.4%) |
|  | <120/<80 and taking medication | 80 | 0 (0%) | 0 (0%) | 0 (0%) | 0 (0%) | 0 (0%) |
|  | <120/< 80 | 100 | 0 (0%) | 0 (0%) | 0 (0%) | 0 (0%) | 0 (0%) |

**Supplemental Table 3.** Life’s Essential 8 points assigned to categories of the Mediterranean Adherence Screener questionnaire.

| **LE8 score (points)** | **MEPA score (points)** | **MEDAS score (points)** |
| --- | --- | --- |
| 100 | 15-16 | 13-14 |
| 80 | 12-14 | 10-12 |
| 50 | 8-11 | 7-9 |
| 25 | 4-7 | 3-6 |
| 0 | 0-3 | 0-2 |

Abbreviations: LE8, Life's Essential 8; MEDAS, Mediterranean Adherence Screener; MEPA, Mediterranean Eating Pattern for Americans;

**Supplemental Table 4.** Descriptive characteristics of the study sample by age groups.

|  | All<br>(n=102) | Adults<br>(n=65) | Older adults<br>(n=37) | *P*-value |
| --- | --- | --- | --- | --- |
| **Demographics** |  |  |  |  |
| Sex, females | 21 (20.6%) | 13 (20%) | 8 (21.6%) | .99 |
| Age, years | 62.3 (6.6) | 58.2 (4.2) | 69.5 (2.8) | **<0.001** |
| Body mass, kg | 83.1 (13.1) | 84.5 (12.6) | 80.7 (13.8) | .17 |
| Height, cm | 168.9 (8.9) | 169.0 (9.0) | 168.7 (8.8) | .85 |
| Education, categories |  |  |  | .44 |
| - Primary school | 20 (19.6%) | 11 (16.9%) | 9 (24.3%) |  |
| - Secondary school | 41 (40.2%) | 29 (44.6%) | 12 (32.4%) |  |
| - University | 41 (40.2%) | 25 (38.5%) | 16 (43.2%) |  |
| **Cardiovascular health-related factors** |  |  |  |  |
| Smoking, categories |  |  |  | .66 |
| - Current smoker | 12 (11.9%) | 9 (13.8%) | 3 (8.1%) |  |
| - Former smoker | 75 (74.2%) | 47 (72.3%) | 28 (75.7%) |  |
| - Never smoker | 14 (13.9%) | 8 (12.3%) | 6 (16.2%) |  |
| MEDAS score (0-14 points) | 7.5 (1.9) | 7.2 (1.8) | 8.0 (2.0) | .06 |
| MVPA, min/week | 358.8 (273.5) | 367.7 (225.2) | 343.1 (345.4) | .15 |
| Sleep duration, h | 7.7 (1.0) | 7.6 (0.9) | 7.9 (1.1) | .18 |
| Body mass index, kg/m<sup>2</sup> | 29.1 (3.8) | 29.6 (3.8) | 28.3 (3.9) | .12 |
| Body mass index, categories |  |  |  | .46 |
| - Healthy weight | 9 (8.8%) | 4 (6.2%) | 5 (13.5%) |  |
| - Overweight | 57 (55.9%) | 35 (53.8%) | 22 (59.5%) |  |
| - Obesity class I | 27 (26.5%) | 19 (29.2%) | 8 (21.6%) |  |
| - Obesity class II | 9 (8.8%) | 7 (10.8%) | 2 (5.4%) |  |
| Diabetes mellitus type II | 18 (17.6%) | 13 (20.0%) | 5 (13.5%) | .58 |
| Fasting blood glucose, mg/dL | 102.9 (29.1) | 102.4 (24.5) | 103.8 (36.1) | .51 |
| HbA1c, % | 5.8 (0.8) | 5.9 (0.8) | 5.8 (0.9) | .65 |
| Non-HDL cholesterol, mg/dL | 88.4 (32.8) | 83.9 (28.5) | 105.8 (41.9) | **.04** |
| Systolic BP, mmHg | 126.9 (14.5) | 126.3 (14.7) | 127.9 (14.3) | .59 |
| Diastolic BP, mmHg | 78.3 (9.0) | 79.2 (9.3) | 76.8 (8.4) | .21 |
| Anti-hypertensive medication | 83 (81.4%) | 54 (83.1%) | 29 (78.4%) | .75 |
| Lipid-lowering medication | 101 (99.0%) | 64 (98.5%) | 37 (100%) | .99 |
| Betablockers | 77 (75.5%) | 50 (76.9%) | 27 (73.0%) | .84 |
| **LE8 score** (0-100 points) | 66.6 (8.6) | 65.6 (8.8) | 68.4 (8.2) | .12 |

|  | All<br>(n=102) | Adults<br>(n=65) | Older adults<br>(n=37) | P-value |
| --- | --- | --- | --- | --- |
| **Health behaviors score**, range 0-100 points | 73.5 (10.3) | 72.5 (10.6) | 75.2 (9.7) | .16 |
| **Health factors score**, range 0-100 points | 59.6 (14.1) | 58.7 (13.6) | 61.2 (14.9) | .39 |
| **Cardiopulmonary exercise test** |  |  |  |  |
| VO<sub>2peak</sub>, mL/kg/min | 25.3 (5.1) | 26.2 (4.7) | 23.7 (5.3) | **.01** |
| VO<sub>2peak</sub>, mL/kg/min, % pred | 93.6 (16.6) | 93.2 (15.5) | 94.3 (18.4) | .96 |
| Time to exhaustion, s | 661.8 (128.7) | 688.8 (119.8) | 614.2 (131.6) | **.004** |
| HR<sub>peak</sub>, beats/min | 138.2 (17.0) | 142.1 (15.6) | 131.4 (17.4) | **.001** |
| HR<sub>peak</sub>, % pred HR<sub>max</sub> | 105.7 (15.5) | 107.0 (15.0) | 103.6 (16.4) | .29 |
| HR<sub>peak</sub>, ≥ 85% pred HR<sub>max</sub> | 94 (92.2%) | 61 (93.8%) | 33 (89.2%) | .46 |
| RER<sub>peak</sub> | 1.183 (0.1) | 1.2 (0.1) | 1.154 (0.1) | **.02** |
| RER<sub>peak</sub> ≥ 1.10, n (%) | 77 (75.5%) | 51 (78.5%) | 26 (70.3%) | .15 |
| RPE<sub>peak</sub>, range 0-10 | 7.7 (1.3) | 7.7 (1.4) | 7.6 (1.3) | .52 |
| HR<sub>rec</sub>, beats/min | 21.6 (8.1) | 23.5 (7.6) | 18.5 (8.1) | **.003** |
| VAT, mL/kg/min of VO<sub>2</sub> | 16.8 (3.4) | 17.2 (3.2) | 16.1 (3.5) | .12 |
| OUES, mL/min/log(L/min) | 765.6 (222.7) | 786.4 (224.4) | 729.1 (217.9) | .21 |
| VO<sub>2</sub> pulse, mL/beat | 15.2 (3.1) | 15.6 (3.1) | 14.5 (3.1) | .08 |
| Systolic BP<sub>peak</sub>, mmHg | 183.9 (23.8) | 184.9 (22.5) | 182.0 (26.3) | .57 |

Data is presented as mean (SD) for continuous variables and as n (%) for categorical variables. Due to missing data, smoking status: n=101 (adults=64, older adults=37), non-HDL cholesterol: n=96 (adults=63, older adults=33), HbA1c: n=98 (adults=62, older adults=36), VO<sub>2peak</sub>: n=100 (adults=63, older adults=37), VO<sub>2peak</sub> (%pred): n=100 (adults=63, older adults=37), RER<sub>peak</sub>: n=95 (adults=59, older adults=36), maximal test (RER<sub>peak</sub> ≥ 1.10): n=95 (adults=59, older adults=36), RPE<sub>peak</sub>: n=98 (adults=62, older adults=36), HR<sub>rec</sub>: n=99 (adults=62, older adults=37), VAT: n=96 (adults=60, older adults=36), VO<sub>2</sub> pulse: n=100 (adults=63, older adults=37), and SBP<sub>peak</sub>: n=98 (adults=64, older adults=34).
Abbreviations: %pred, percent of predicted; BP, blood pressure; HbA1c, glycosylated hemoglobin; HR, heart rate; HR<sub>rec</sub>, heart rate recovery; LE8, Life's Essential 8; MEDAS, Mediterranean Diet Adherence Screener; MVPA, moderate-to-vigorous intensity physical activity; non-HDL, non-high-density lipoprotein; OUES, oxygen uptake efficiency slope; RER, respiratory exchange ratio; RPE, rating of perceived exertion; VAT, ventilatory anaerobic threshold; VO<sub>2</sub>, oxygen uptake.

**Supplemental Table 5.** Descriptive characteristics of the study sample by education groups.

|  | All<br>(n=102) | No university<br>(n=61) | University<br>(n=41) | *P*-value |
| --- | --- | --- | --- | --- |
| **Demographics** |  |  |  |  |
| Sex, females | 21 (20.6%) | 14 (23%) | 7 (17.1%) | .64 |
| Age, years | 62.3 (6.6) | 62.2 (6.5) | 62.3 (6.9) | .96 |
| Body mass, kg | 83.1 (13.1) | 83.9 (13.6) | 82.0 (12.3) | .49 |
| Height, cm | 168.9 (8.9) | 168.0 (9.6) | 170.2 (7.6) | .23 |
| **Cardiovascular health-related factors** |  |  |  |  |
| Smoking, categories |  |  |  | .50 |
| - Current smoker | 12 (11.9%) | 9 (14.8%) | 3 (7.3%) |  |
| - Former smoker | 75 (74.2%) | 43 (70.5%) | 32 (78.0%) |  |
| - Never smoker | 14 (13.9%) | 9 (14.8%) | 5 (12.2%) |  |
| MEDAS score (0-14 points) | 7.5 (1.9) | 7.5 (2.0) | 7.5 (1.8) | .77 |
| MVPA, min/week | 358.8 (273.5) | 390.9 (332.3) | 311.0 (138.9) | .72 |
| Sleep duration, h | 7.7 (1.0) | 7.8 (1.0) | 7.6 (1.0) | .19 |
| Body mass index, kg/m<sup>2</sup> | 29.1 (3.8) | 29.6 (3.6) | 28.3 (4.1) | **.02** |
| Body mass index, categories |  |  |  | .32 |
| - Healthy weight | 9 (8.8%) | 4 (6.6%) | 5 (12.2%) |  |
| - Overweight | 57 (55.9%) | 32 (52.5%) | 25 (61.0%) |  |
| - Obesity class I | 27 (26.5%) | 20 (32.8%) | 7 (17.1%) |  |
| - Obesity class II | 9 (8.8%) | 5 (8.2%) | 4 (9.8%) |  |
| Diabetes mellitus type II | 18 (17.6%) | 12 (19.7%) | 6 (14.6%) | .70 |
| Fasting blood glucose, mg/dL | 102.9 (29.1) | 104.1 (33.1) | 101.0 (21.9) | .93 |
| HbA1c, % | 5.8 (0.8) | 5.9 (1.0) | 5.7 (0.5) | .72 |
| Non-HDL cholesterol, mg/dL | 88.4 (32.8) | 94.3 (37.2) | 79.6 (22.2) | .07 |
| Systolic BP, mmHg | 126.9 (14.5) | 128.9 (14.5) | 123.8 (14.1) | .08 |
| Diastolic BP, mmHg | 78.3 (9.0) | 79.7 (9.4) | 76.2 (8.1) | .05 |
| Anti-hypertensive medication | 83 (81.4%) | 53 (86.9%) | 30 (73.2%) | .14 |
| Lipid-lowering medication | 101 (99.0%) | 60 (98.4%) | 41 (100%) | .99 |
| Betablockers | 77 (75.5%) | 45 (73.8%) | 32 (78.0%) | .80 |
| **LE8 score** (0-100 points) | 66.6 (8.6) | 65.0 (8.9) | 69.1 (7.7) | **.02** |
| **Health behaviors score**, range 0-100 points | 73.5 (10.3) | 72.5 (11.3) | 74.9 (8.6) | .41 |
| **Health factors score**, range 0-100 points | 59.6 (14.1) | 57.2 (14.9) | 63.3 (12.0) | **.03** |

|  | All<br>(n=102) | No university<br>(n=61) | University<br>(n=41) | P-value |
| --- | --- | --- | --- | --- |
| **Cardiopulmonary exercise test** |  |  |  |  |
| VO<sub>2peak</sub>, mL/kg/min | 25.3 (5.1) | 24.9 (5.2) | 25.9 (4.9) | .31 |
| VO<sub>2peak</sub>, mL/kg/min, % pred | 93.6 (16.6) | 95.2 (17.4) | 91.4 (15.2) | .25 |
| Time to exhaustion, s | 661.8 (128.7) | 653.7 (138.3) | 673.7 (113.5) | .44 |
| HR<sub>peak</sub>, beats/min | 138.2 (17.0) | 137.5 (16.8) | 139.4 (17.4) | .59 |
| HR<sub>peak</sub>, % pred HR<sub>max</sub> | 105.7 (15.5) | 104.3 (14.4) | 107.9 (17.0) | .12 |
| HR<sub>peak</sub>, ≥ 85% pred HR<sub>max</sub> | 94 (92.2%) | 56 (91.8%) | 38 (92.7%) | .99 |
| RER<sub>peak</sub> | 1.183 (0.1) | 1.166 (0.1) | 1.208 (0.1) | **.04** |
| RER<sub>peak</sub> ≥ 1.10, n (%) | 77 (75.5%) | 42 (68.9%) | 35 (85.4%) | **.03** |
| RPE<sub>peak</sub>, range 0-10 | 7.7 (1.3) | 7.6 (1.4) | 7.7 (1.3) | .83 |
| HR<sub>rec</sub>, beats/min | 21.6 (8.1) | 20.9 (8.6) | 22.6 (7.4) | .32 |
| VAT, mL/kg/min of VO<sub>2</sub> | 16.8 (3.4) | 16.7 (3.5) | 17.0 (3.2) | .71 |
| OUES, mL/min/log(L/min) | 765.6 (222.7) | 758.1 (234.3) | 776.8 (206.6) | .68 |
| VO<sub>2</sub> pulse, mL/beat | 15.2 (3.1) | 15.2 (3.3) | 15.3 (2.8) | .84 |
| Systolic BP<sub>peak</sub>, mmHg | 183.9 (23.8) | 182.4 (25.3) | 186.0 (21.6) | .47 |

Data is presented as mean (SD) for continuous variables and as n (%) for categorical variables. Due to missing data, smoking status: n=101 (no university=61, university=40), non-HDL cholesterol: n=96 (no university=58, university=38), HbA1c: n=98 (no university=57, university=41), VO<sub>2peak</sub>: n=100 (no university=59, university=41), VO<sub>2peak</sub> (%pred): n=100 (no university=59, university=41), RER<sub>peak</sub>: n=95 (no university=57, university=38), maximal test (RER<sub>peak</sub> ≥1.10): n=95 (no university=57, university=38), RPE<sub>peak</sub>: n=98 (no university=58, university=40), HR<sub>rec</sub>: n=99 (no university=58, university=41), VAT: n=96 (no university=58, university=38), VO<sub>2</sub> pulse: n=100 (no university=59, university=41), and SBP<sub>peak</sub>: n=98 (no university=57, university=41).
Abbreviations: %pred, percent of predicted; BP, blood pressure; HbA1c, glycosylated hemoglobin; HR, heart rate; HR<sub>rec</sub>, heart rate recovery; LE8, Life's Essential 8; MEDAS, Mediterranean Diet Adherence Screener; MVPA, moderate-to-vigorous intensity physical activity; non-HDL, non-high-density lipoprotein; OUES, oxygen uptake efficiency slope; RER, respiratory exchange ratio; RPE, rating of perceived exertion; VAT, ventilatory anaerobic threshold; VO<sub>2</sub>, oxygen uptake.

**Supplemental Table 6.** Unadjusted Pearson correlations *P*-values (See correlation coefficients in Figure 2).

|  | Sex | Age | Education | Time to exhaustion | VO<sub>2peak</sub><br>(mL/kg/min) | VAT<br>(mL/kg/min) | Oxygen pulse | Heart rate recovery | OUES | Health behaviors | Health factors |
| --- | --- | --- | --- | --- | --- | --- | --- | --- | --- | --- | --- |
| **Age** | .77 |  |  |  |  |  |  |  |  |  |  |
| **Education** | .16 | .68 |  |  |  |  |  |  |  |  |  |
| **Time to exhaustion** | <.001 | <.001 | .24 |  |  |  |  |  |  |  |  |
| **VO<sub>2peak</sub>**<br>**(mL/kg/min)** | <.001 | <.001 | .21 | <.001 |  |  |  |  |  |  |  |
| **VAT**<br>**(mL/kg/min)** | .004 | .06 | .48 | <.001 | <.001 |  |  |  |  |  |  |
| **Oxygen pulse** | <.001 | .09 | .79 | <.001 | <.001 | <.001 |  |  |  |  |  |
| **Heart rate recovery** | .34 | .008 | .42 | .002 | <.001 | <.001 | .38 |  |  |  |  |
| **OUES** | <.001 | .05 | .96 | <.001 | <.001 | <.001 | <.001 | .94 |  |  |  |
| **Health behaviors** | .23 | .37 | .65 | .01 | .05 | .08 | .58 | .71 | .80 |  |  |
| **Health factors** | .81 | .62 | .02 | <.001 | <.001 | .003 | .15 | .005 | .26 | .99 |  |
| **LE8 total score** | .63 | .30 | .03 | <.001 | <.001 | <.001 | .43 | .02 | .30 | <.001 | <.001 |
| Abbreviations: OUES, oxygen uptake efficiency slope; VAT, ventilatory anaerobic threshold; VO<sub>2peak</sub>, peak oxygen uptake. |  |  |  |  |  |  |  |  |  |  |  |

**Supplemental Table 7.** Linear regressions between cardiorespiratory fitness indicators and Life’s Essential 8 scores.

| CPET indicators | Life's Essential 8 |  |  | Health behaviors |  |  | Health factors |  |  |
| --- | --- | --- | --- | --- | --- | --- | --- | --- | --- |
|  | B | 95% CI | *P*-value | B | 95% CI | *P*-value | B | 95% CI | *P*-value |
| Time to exhaustion (s) | 0.043 | 0.030; 0.055 | **<.001** | 0.035 | 0.018; 0.052 | **<.001** | 0.051 | 0.029; 0.074 | **<.001** |
| VO<sub>2peak</sub> (mL/kg/min) | 1.14 | 0.83; 1.45 | **<.001** | 0.74 | 0.29; 1.19 | **.002** | 1.57 | 1.02; 2.13 | **<.001** |
| VAT (mL/kg/min) | 1.10 | 0.59; 1.61 | **<.001** | 0.84 | 0.19; 1.49 | **.01** | 1.45 | 0.58; 2.32 | **.001** |
| OUES | -0.003 | -0.01; 0.007 | .54 | 0.003 | -0.009; 0.01 | .61 | -0.009 | -0.02; 0.006 | .23 |
| Oxygen pulse (mL/beat) | -0.14 | -0.93; 0.65 | .73 | 1.07 | 0.13; 2.006 | **.02** | -1.40 | -2.67; -0.13 | **.03** |
| Heart rate recovery (beats per minute) | 0.27 | 0.06; 0.49 | **.01** | 0.05 | -0.21; 0.31 | .71 | 0.52 | 0.16; 0.87 | **.004** |
| Abbreviations: B, unstandardized regression coefficient; OUES, oxygen uptake efficiency slope; VAT, ventilatory anaerobic threshold; VO<sub>2peak</sub>, peak oxygen uptake. |  |  |  |  |  |  |  |  |  |
| The results represent <u>unstandardized</u> regression coefficients from linear regression analyses, with 95% confidence intervals and *P*-values. All analyses were adjusted for sex, age and education. Significant *P*-values are presented in bold. |  |  |  |  |  |  |  |  |  |

**Supplemental Table 8.** Sensitivity analyses of linear regressions between cardiopulmonary exercise test indicators and Life’s Essential 8 scores in tests in which peak respiratory exchange ratio was ≥1.10 and peak heart rate was ≥85% of predicted peak heart rate (n=76).

| CPET indicators | Life's Essential 8 |  |  | Health behaviors |  |  | Health factors |  |  |
| --- | --- | --- | --- | --- | --- | --- | --- | --- | --- |
| | $\beta_{\text{std}}$ | 95% CI | *P*-value | $\beta_{\text{std}}$ | 95% CI | *P*-value | $\beta_{\text{std}}$ | 95% CI | *P*-value |
| **Time to exhaustion** | 0.54 | 0.31; 0.78 | **<.001** | 0.36 | 0.10; 0.62 | **.007** | 0.38 | 0.12; 0.64 | **.004** |
| **VO<sub>2peak</sub> (mL/kg/min)** | 0.63 | 0.43; 0.84 | **<.001** | 0.34 | 0.10; 0.59 | **.007** | 0.50 | 0.27; 0.73 | **<.001** |
| **VAT (mL/kg/min)** | 0.42 | 0.21; 0.63 | **<.001** | 0.31 | 0.08; 0.54 | **.009** | 0.28 | 0.05; 0.51 | **.02** |
| **OUES** | -0.13 | -0.40; 0.13 | .32 | 0.05 | -0.22; 0.32 | .72 | -0.21 | -0.48; 0.06 | .12 |
| **Oxygen pulse** | -0.06 | -0.39; 0.27 | .73 | 0.22 | -0.11; 0.54 | .19 | -0.23 | -0.56; 0.09 | .16 |
| **Heart rate recovery** | 0.13 | -0.11; 0.37 | .29 | 0.01 | -0.23; 0.26 | .90 | 0.15 | -0.10; 0.40 | .25 |
| Abbreviations: OUES, oxygen uptake efficiency slope; VAT, ventilatory anaerobic threshold; VO<sub>2peak</sub>, peak oxygen uptake. |  |  |  |  |  |  |  |  |  |
| The results represent standardized beta coefficients from linear regression analyses, with 95% confidence intervals and *P*-values. All analyses were adjusted for sex, age, and education. Significant *P*-values are presented in bold. |  |  |  |  |  |  |  |  |  |

